# Integrative analyses identify susceptibility genes underlying COVID-19 hospitalization

**DOI:** 10.1101/2020.12.07.20245308

**Authors:** Gita A Pathak, Kritika Singh, Tyne W Miller-Fleming, Frank R Wendt, Nava Ehsan, Kangcheng Hou, Ruth Johnson, Zeyun Lu, Shyamalika Gopalan, Loic Yengo, Pejman Mohammadi, Bogdan Pasaniuc, Renato Polimanti, Lea K Davis, Nicholas Mancuso

## Abstract

Despite rapid progress in characterizing the role of host genetics in SARS-Cov-2 infection, there is limited understanding of genes and pathways that contribute to COVID-19. Here, we integrated a genome-wide association study of COVID-19 hospitalization (7,885 cases and 961,804 controls from COVID-19 Host Genetics Initiative) with mRNA expression, splicing, and protein levels (n=18,502). We identified 27 genes related to inflammation and coagulation pathways whose genetically predicted expression was associated with COVID-19 hospitalization. We functionally characterized the 27 genes using phenome- and laboratory-wide association scans in Vanderbilt Biobank (BioVU; n=85,460) and identified coagulation-related clinical symptoms, immunologic, and blood-cell-related biomarkers. We replicated these findings across trans-ethnic studies and observed consistent effects in individuals of diverse ancestral backgrounds in BioVU, pan-UK Biobank, and Biobank Japan. Our study highlights putative causal genes impacting COVID-19 severity and symptomology through the host inflammatory response.

**SINGLE-SENTENCE SUMMARY:** Large-scale genomic studies identify genes in the inflammation and coagulation pathways contributing to risk and symptomology of COVID-19 disease.

---

Coronavirus disease 2019 (COVID-19), caused by the severe acute respiratory syndrome coronavirus 2 (SARS-CoV-2), was first reported in December 2019 and rapidly progressed into a global pandemic (*1*). Approximately 10-20% of patients known to be infected with the respiratory virus SARS-CoV-2 need hospitalization (*2*), and among them, a fraction face significant morbidity and mortality (*3*). The host’s genetic background is likely to contribute in explaining such diverse clinical outcomes. While previous efforts have demonstrated the role of *ACE2* and *TMPRSS2* in host defense against COVID-19 (*4*), there remains limited understanding for the role of host genetics contributing to severe COVID-19 outcome variability.

Here we integrated mRNA expression, splicing, and protein abundance data (n=18,502) with GWAS of COVID-19 related hospitalization (n=7,885 cases, 961,804 controls; Freeze 4 COVID-19 HGI excluding 23andMe participants (*5*–*9*)) to map genes and pathways involved in COVID-19 severity. We performed mRNA/splicing/protein transcriptome-wide association studies (TWAS/spTWAS/PWAS) to identify 27 genes across 13 genomic regions whose genetically predicted activity is associated with COVID-19 related hospitalization. We investigated the functional role of these 27 genes using phenome-wide (PheWAS) and laboratory-wide (LabWAS) association scans to map their role in immunity and blood biomarkers in European and African ancestry patients from the Vanderbilt University Medical Center biobank (BioVU; n=85,460). We replicated phenotypes identified from BioVU in secondary cohorts of multi-ethnic individuals from the Pan-UK Biobank (980 Admixed American, 6,636 African, 8,876 Central/South Asian, 2,709 East Asian, 420,531 European, and 1,599 Middle Eastern) and Biobank Japan (up to 212,453 Japanese participants). Taken together, our results suggest multiple molecular mechanisms contributing to severe COVID-19 outcomes and highlight potential therapeutic targets.

## RESULTS

### TWAS identifies genes for COVID-19 related hospitalization

To identify genes underlying COVID-19 related hospitalization, we tested the predicted expression of 22,207 genes across 49 tissues for association with COVID-19 related hospitalization (*see Methods*). We identified 123 associations representing 21 genes across 45 tissues at 8 independent genomic regions (p-value < 2.3E-6; **Fig. 1, 2**; **Tables S1-S3**). Next, to improve statistical power, we tested for association between predicted gene expression levels from multiple tissues simultaneously with COVID-19 related hospitalization GWAS. Of the 22,207 tested genes, we identified 14 genes across 10 genomic regions, which consisted of 2 additional genes – *XCR1* and *DNAH3* (p-value < 1.4E-06; **Fig. S1.; Table S4**.). Overall, we found 23 TWAS-based gene associations across 10 genomic regions.

**Figure 1:**
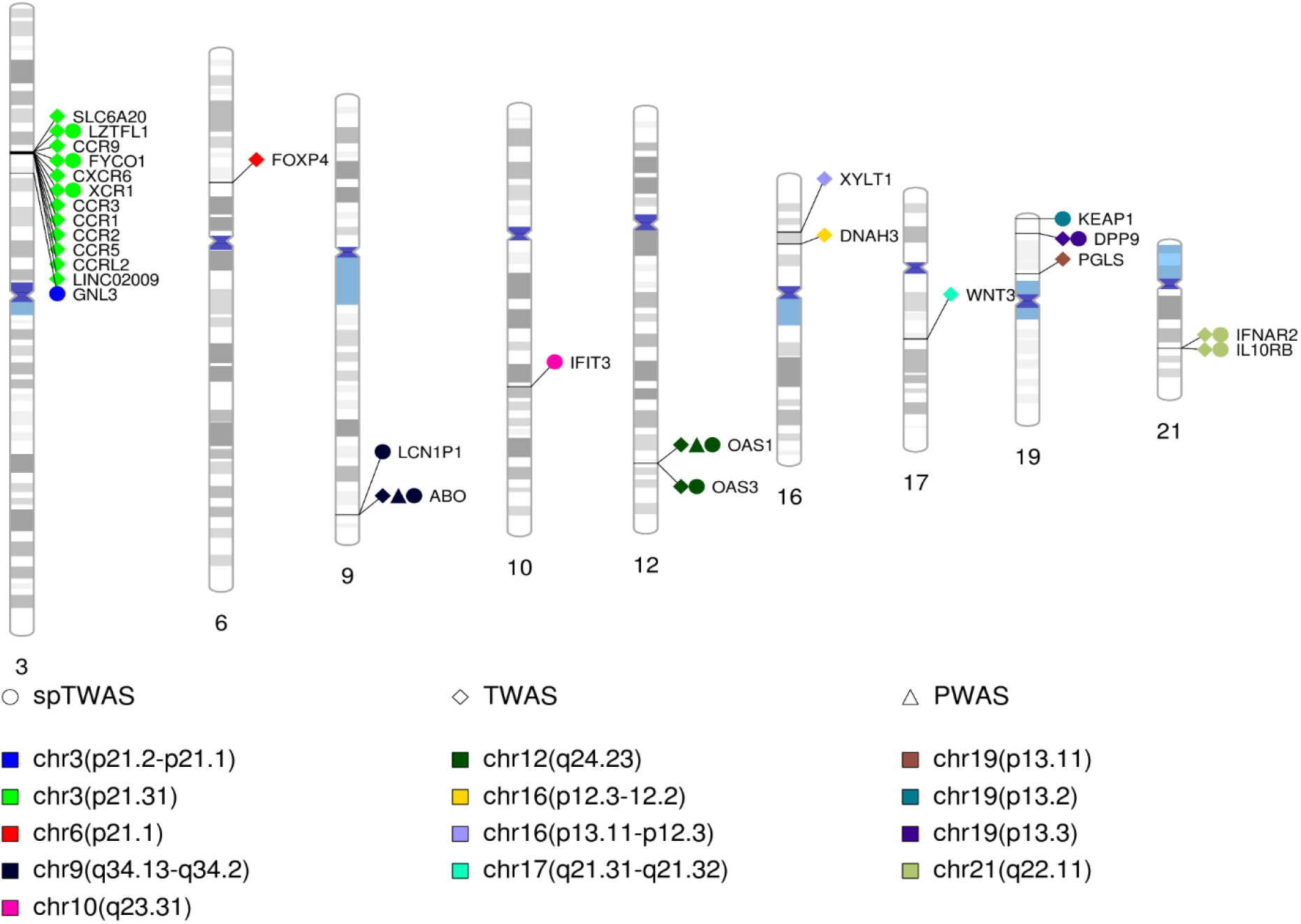
Significant genes. The integrative analyses identified 27 genes (labeled) across 13 regions color coded) shown in the ideogram.

**Figure 2:**
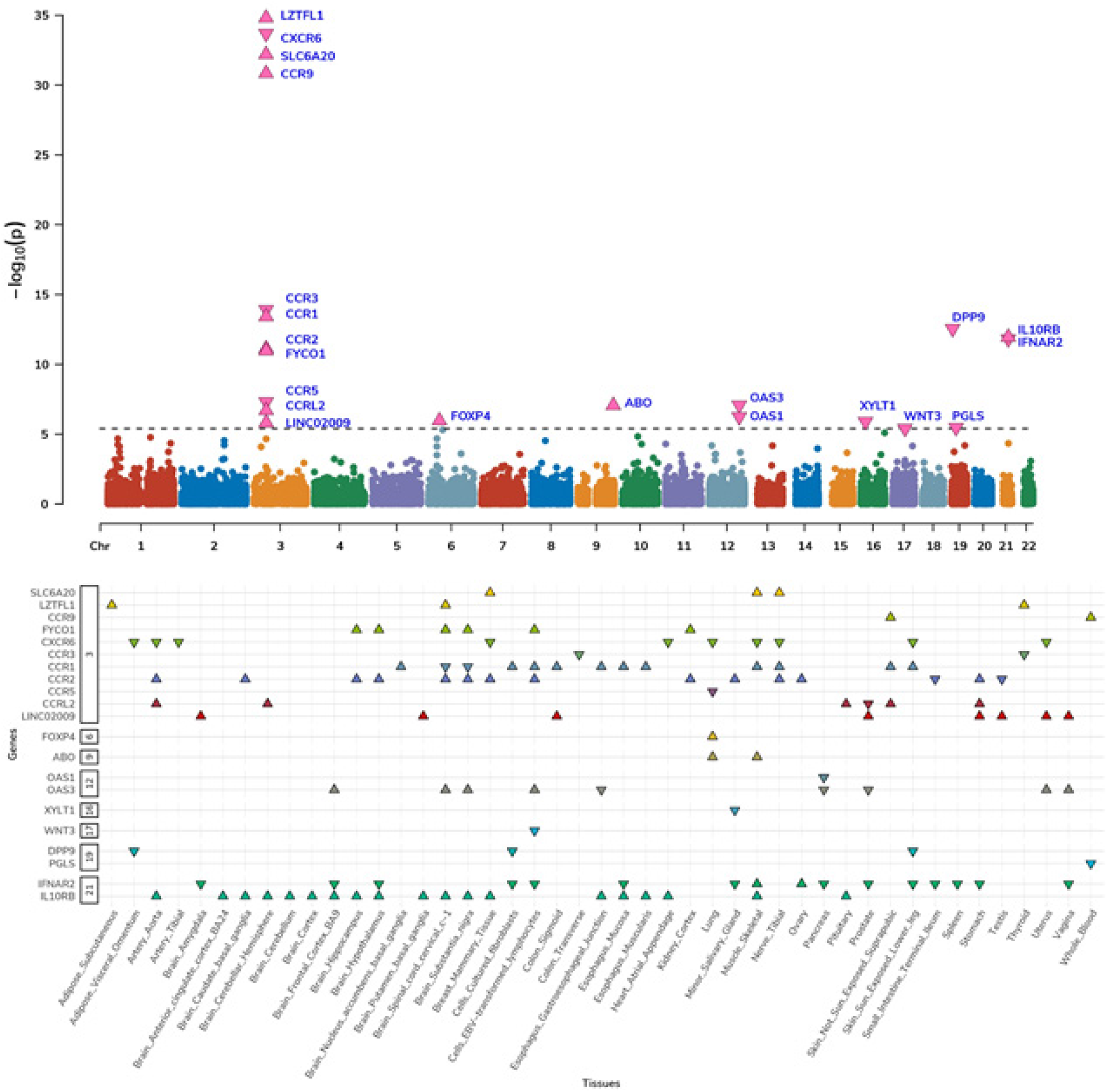
TWAS: The top panel is a Manhattan plot of genes associated via multiple-tissue TWAS. Each data point represents a gene grouped by chromosome (x-axis) and lowest p-value (y-axis) of the gene across significant tissues. The significant genes are shown as pink triangles, wherein triangles facing up and down represent positive and negative z-scores, respectively. The bottom panel show distribution of z-scores across significant gene-tissue airs. The genes are grouped based on chromosome (y-axis) and respective tissues (x-axis).

To find additional support for genetic regulation of identified susceptibility genes by SNPs at risk regions, we tested the 123 gene/tissue pairs identified in single-tissue scans for allelic imbalance within 1Mb GWAS regions (see Methods). We identified 9 genes (*ABO, CCR2, CXCR6, FYCO1, IFNAR2, IL10RB, LZTFL1, OAS1, OAS3*) with evidence of allelic imbalance at COVID-19 GWAS risk variants, with 3 genes (*ABO, OAS3, IL10RB*; see **Fig. S2**., **Table S5**.) when restricted to leading GWAS index variants (p-value < 0.05 / 21). Together, these results further support a model where risk is conferred through genetic dysregulation at susceptibility genes.

Next, we focused on the impact of genetic regulation of alternative splicing for COVID-19 severity and performed a multi-tissue splicing transcriptome-wide association study (spTWAS; see Methods). Overall, we tested 131,376 splice sites of predicted alternative-splicing expression across 49 tissues for association with COVID19 related hospitalization and identified 420 associations representing 43 splice variants for 11 genes across 49 tissues and 5 genomic regions (see **Fig. 1, 3**.; **Tables S6.-S8**.). Next, we performed a multi-tissue analysis (see Methods) and identified 34 splice variants for 12 genes (2 genes – *IFIT3* and *GNL3* not identified in single-tissue scans) across 40 tissues (p-value < 3.7E-07; see **Fig. S3**.; **Table S9**.).

**Figure 3:**
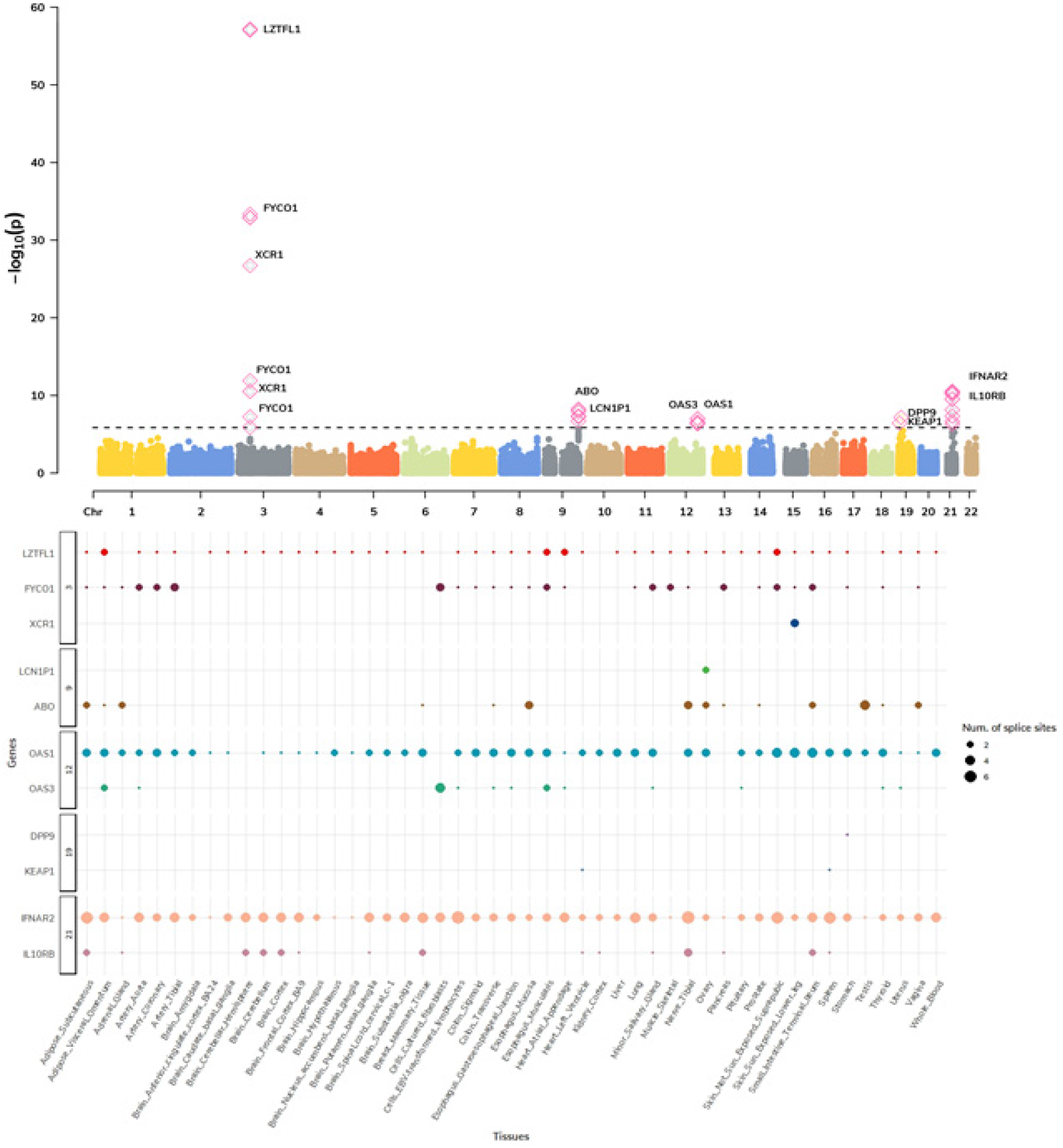
Splicing TWAS. The top panel is a Manhattan plot of genes associated via multiple tissue spTWAS. Each data point represents splice site grouped by chromosome (x-axis) and lowest p-value (y-axis) of the splice site across significant tissues. The annotated genes to splice site are labeled. The significant splice sites are shown as pink diamonds. The bottom panel show distribution of splice sites across significant site-tissue pairs. The genes annotated to splice sites are grouped based on chromosome (y-axis) and respective tissues (x-axis).

Comparing genes identified from TWAS (23 genes) and spTWAS (13 genes), 9 genes were implicated by both approaches - *LZTFL1, DPP9, IL10RB, IFNAR2, OAS3, FYCO1, ABO, OAS1* and *XCR1*. Alternative splicing had stronger overall association signals at the 9 genes in common (p-value = 2.17E-09), with 5/9 genes showing greater signals on average (p-value < 0.05 / 9; see T**able S10**.).

Next, we interrogated the role of genetic regulation of protein abundances and performed a proteome-wide association study (PWAS) using 1,031 predictive models of plasma proteins fitted from population data in the INTERVAL study (N=3,301; see Methods) (*10*). Of the 1,031 tests performed, 2 genes (*ABO*, and *OAS1*) were significantly associated with COVID-19 related hospitalization (p-value < 4.85E-5; **Fig. 1, Table S11**.).

### Phenome- and laboratory-wide association scans highlight functional role for the 27 genes

We investigated the potential functional role of the 27 TWAS genes using data of 1,404 clinical phenotypes for N=70,439 individuals of European ancestry using the Vanderbilt Biobank, BioVU (see Methods; **Fig. 4**., **Table S12**.). Overall, 40 clinical phenotypes were significantly associated with genetically-predicted *ABO, IFNAR2*, and *CCR1* expression levels; *ABO* accounted for the majority (30 out of 40) of the associations. Across the 17 phenotype categories, we found circulatory system-related phenotypes were enriched for association (see Methods; 7.23-fold enrichment, p-value = 8.62E-22, **Table S13**). We repeated an enrichment analysis using all association data, regardless of statistical significance, and observed circulatory- and infectious disease-related phenotypes strongly enriched for association signal on average (p-value < 3.9E-44; **Table S14**). Top associations with genetically-derived *ABO* gene expression were driven by circulatory system phenotypes, including acute pulmonary heart disease, deep vein thrombosis, other venous embolism and thrombosis, pulmonary heart disease, and acute pulmonary thrombosis and infarction (OR=1.47; p-value = 3.97E-11). *IFNAR2* was associated with migraine (OR = 1.35; p-value = 4.10E-06) and with throat pain (OR = 2.05; p-value = 2.62E-05; **Table S12**).

**Figure 4:**
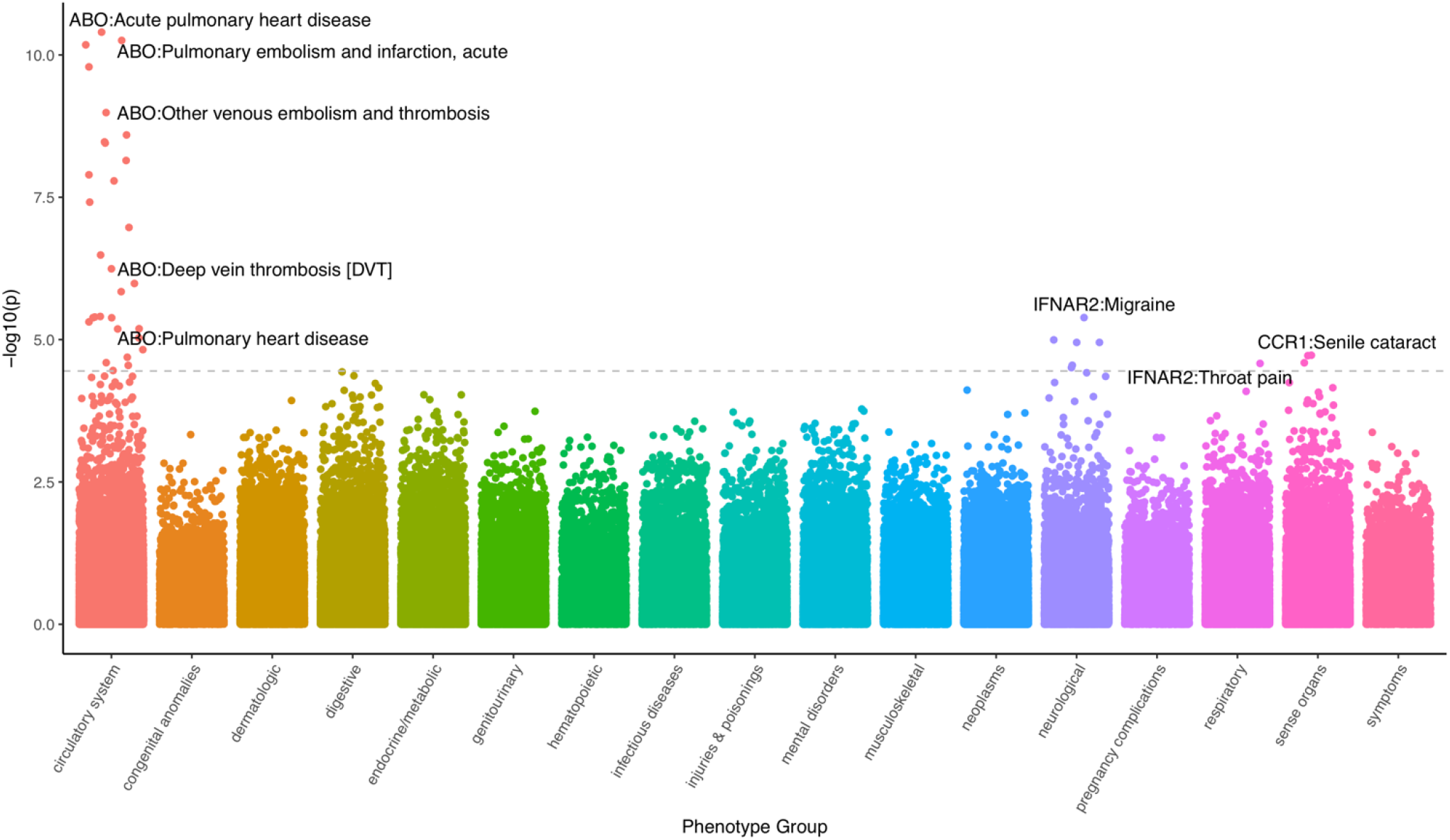
PheWAS Manhattan Plot. Each data point represents phenotypic associations with genetically-regulated expression of gene-tissue pairs. The data points are grouped and color-coded by phenotype groups x-axis) and -log10(p-value) (y-axis). The dashed line represents Bonferroni threshold, and most significant gene-phenotype associations across all significant tissues are text-labeled.

Next, we focused on laboratory results for N=70,337 individuals of European ancestry using the Vanderbilt Biobank, BioVU (see Methods). For the 323 laboratory traits tested, we found 32 labs significantly associated with four genes (*ABO, IFNAR2, KEAP1, and SLC6A20*; p-value < 1.55E-04; see **Fig. 5**., **Table S15**.). Of these, *ABO* captured 27/32 significant associations (mean OR = 1.33; p-value = 8.02E-14 < 1.40E-04). Across the 12 broad lab definitions in our data, our significant LabWAS findings were enriched for blood-related lab measurements (see Methods; 4.21-fold enrichment, p-value = 1.23E-11; **Table S16**.). When extending our enrichment analyses to all associations, we found blood-(see Methods; p-value = 9.23E-22; **Table S17**.) and immune-related labs (p-value = 2.81E-14) displayed the greatest enrichment, with toxicology-, urinary-, and cancer-related labs exhibiting significant depletion of signal (**Table S17**.). Genetically-predicted *ABO* expression was associated with various measures of blood and platelet count, coagulation factors, and ferritin in blood, as well as labs measuring immune and metabolic function (T**able S15**.). Genetically-predicted *IFNAR2* expression was negatively associated with creatine kinase (OR=0.89; p=5.85E-05; **Table S15**.). Genetically-predicted *KEAP1* expression was positively associated with total cholesterol, and non-high-density lipoprotein levels (beta= 0.50, p=3.72E-05). *SLC6A20* genetically-predicted expression was negatively associated with basophil volume in blood (beta = -0.19, p= 4.04e-5) and magnesium volume in serum/plasma (beta= -0.26, p= 1.04e-4).

**Figure 5:**
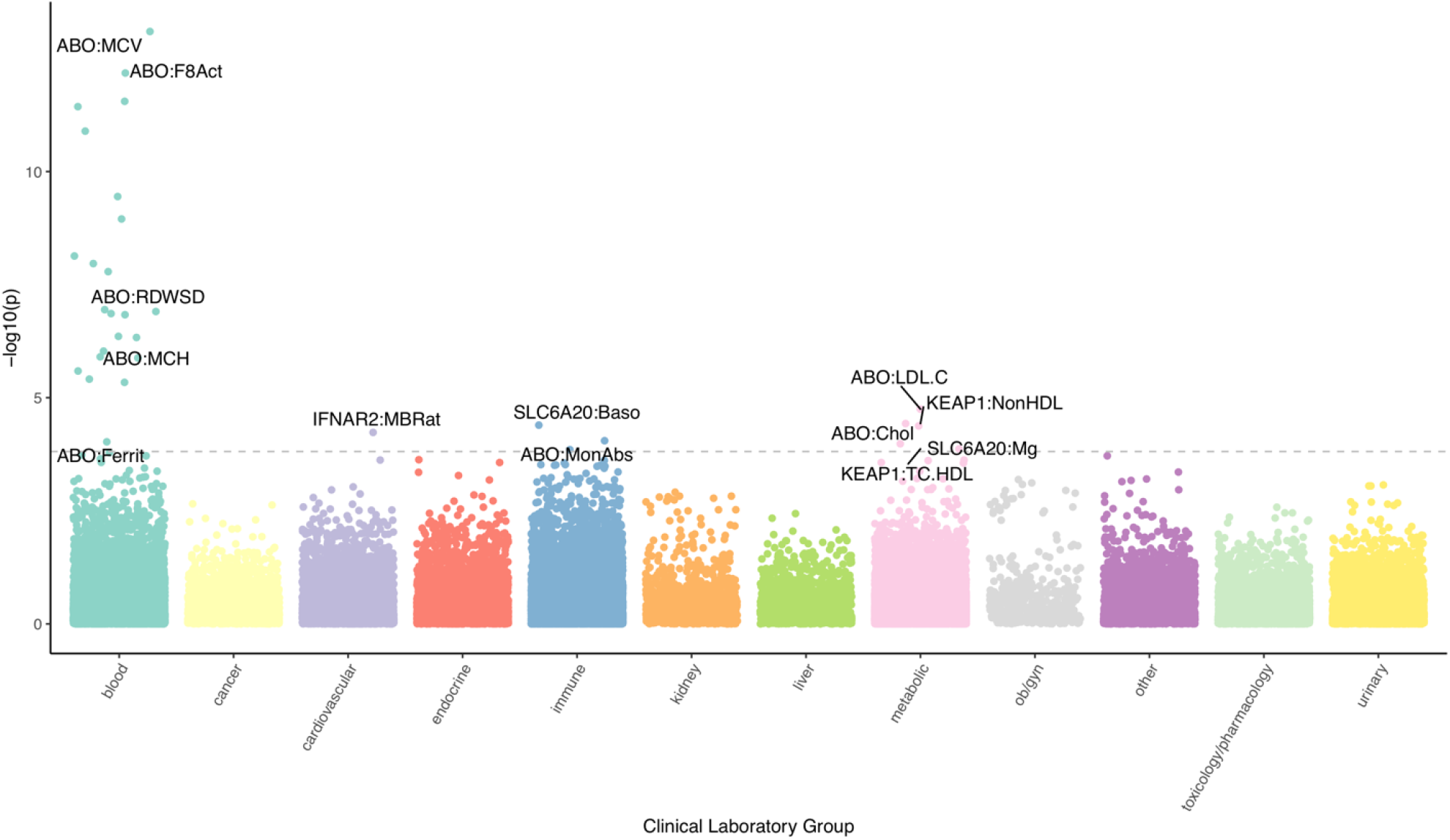
LabWAS Manhattan Plot. Each data point represents laboratory-trait associations with genetically-regulated expression of gene-tissue pairs. The data points are grouped and color-coded by clinical laboratory-test groups (x-axis) and -log10(p-value) (y-axis). The dashed line represents Bonferroni threshold, and most significant gene-laboratory trait associations across all significant tissues are text-labeled.

### Cross-ancestry phenotypic comparisons

To perform cross-ancestry validation of the clinical and laboratory phenotypes implicated in the European-based results, we performed PheWAS and LabWAS in the N=15,123 individuals of African ancestry in the BioVU records. Of the 32 identified laboratory measures, we found 22 *ABO-*associated labs replicated at nominal levels (p-value < 0.05) with 5 replicating after adjusting for the number of tests performed (p-value < 0.05 / 32). We attribute lack of statistical power to the 17 phenotypes that did not replicate after multiple testing correction. Effect sizes across ancestries were highly concordant (slope=0.87, 95CI [0.73, 1.01], p-value = 1.66-13; see Methods; **Table S15**), with no individual gene/lab pair demonstrating evidence of significant heterogeneity (p-value < 0.05 / 32). Of the 40 clinical phenotypes identified in participants of European ancestry, we found none that replicated at Bonferroni adjusted levels. This was largely due to reduced statistical power from smaller sample sizes, as estimated effect-sizes were similar across ancestries (slope=0.33, 95CI[0.25, 0.41], p-value = 6.72E-10; see Methods**; Table S12**.). These cross-ancestry analyses suggest similar effects of predicted expression on relevant clinical and laboratory phenotypes.

We next sought to test cross-ancestry replication of identified phenotypes in the Pan-UK Biobank and Biobank Japan. Briefly, we performed PheWAS using LD-independent eQTL/sQTL SNPs (see methods) of TWAS-identified genes. We identified 233 FDR significant SNP-phenotype results dominated by the associations between *ABO* (214/233, 91.8%) SNPs and blood differential tests such as basophils and monocytes. A subset of 80/233 (34.3%) FDR significant associations also were nominally significant in at least one population of non-European ancestry (p-value < 0.05; **Tables S18.-S19**.). There were six instances of FDR significant effect estimate heterogeneity across ancestries, all of which involved *ABO* SNPs and the biomarker alkaline phosphatase or erythrocyte properties. We next tested how significant EUR effect estimates reflect SNP effects across ancestries. We found that EUR SNP effects significantly predicted SNP effects in six global ancestry groups (maximum prediction in AMR; **Table S20**.).

## DISCUSSION

COVID-19 disease is characterized by a wide variability in presentations and severity. We integrated multi-tiered regulatory information with publicly available variant-level data to identify genes associated with COVID-19 related hospitalization. To investigate the clinical relevance of these findings, we performed a phenome-wide and lab-wide assessment of the genetically predicted mRNA expression value of each gene that was significantly associated with COVID-19. We further examined these associations across diverse ancestries and found nominal replication of blood cell traits in diverse ancestral cohorts of Pan-UKBB, BBJ, and an African American population in BioVU.

All three TWAS approaches (mRNA expression, splicing, and protein expression) identified two genes – *ABO* at 9q34.2 and *OAS1* at 12q24.13. PheWAS results implicated *ABO* in several thrombotic and coagulation-related phenotypes. Thrombotic complications are reported to be both risk factors and sequalae to COVID-19 diagnosis. For example, coagulopathic conditions such venous thromboembolism (*11, 12*), deep vein thrombosis (*13*), and pulmonary heart disease and embolism (*14, 15*) constitute of more than 30% prevalent disorders in hospitalized COVID-19 patients. Abnormal blood cell indices are a common denominator shared by severe COVID-19 (*16*) and thrombotic disorders (*17*). These phenotypic observations are further supported by lab-trait associations which showed predicted expression of *ABO* was associated with coagulation factor VIII, which is critical for thrombotic homeostasis regulated by its carrier protein - von Willebrand factor (*18, 19*). Analysis in individuals of both European and African ancestry supported predicted *ABO* expression associating with blood differential tests including mean corpuscle volume, monocyte count, and erythrocytes.

We observed that associated phenotypes and gene functions converged on cytokine-cytokine receptor signaling involved in inflammatory response (e.g., *CXCR6, CCR9, CCR5, XCR1, IFNAR2, IL10RB*), and on JAK-STAT signaling pathways involved in antiviral host response (e.g., *IFNAR2, OAS1, OAS3*). *IFNAR2* encodes the interferon-alpha/beta receptor beta chain and is responsible for stimulating interferon response which is critical for anti-viral immunity previously observed in influenza viral infection (*20*). *IFNAR2* is hypothesized to modulate immune response to COVID-19 (*21*–*23*) and interferon deficiency is reportedly associated with severe symptoms of COVID-19 (*24*–*26*). These findings are reinforced by results showing that reduced expression (observed in 14 of the 16 significant tissues) of *IFNAR2* is associated with COVID-19 related hospitalization. Genetically predicted expression of *IFNAR2* was associated with migraine in BioVU patients without severe COVID-19. Given that more than 10% of the COVID-19 diagnosed individuals requiring hospitalization reported migraine and headache symptoms (*27, 28*), we hypothesize that reduced host expression of *IFNAR2* may modulate risk for migraine symptoms in the context of severe COVID-19 infection.

Our results are consistent with previous studies investigating the impact of inflammation on severe COVID-19 outcomes; however, we note there are limitations. First, TWAS analyses rely on SNP-based predictive models of mRNA and alternative splicing trained using mostly European-ancestry individuals in GTEx v8 (*29*). While consistent with the ancestry makeup of COVID-19 HGI GWAS ((*6*); https://www.covid19hg.org/), applying these models to non-European individuals (e.g., African Americans in BioVU) will result in loss of power or bias due to different underlying linkage disequilibrium patterns. Second, TWAS uses mRNA, alternative splicing, or protein levels in bulk tissue, with cell-type effects likely to be missed. Third, TWAS assumes additivity of SNP effects on gene expression and downstream hospitalization risk, which ignores the possibility of epistatic and gene-environment interactions contributing to COVID-19 related hospitalization risk. Finally, our study focuses on the host genetic factors that contribute to severe COVID-19 but did not incorporate the social determinants of health that are known to influence risk for severe COVID-19. The biological insights identified here should not be interpreted as explanatory factors for disparity, but instead as key genomic pathways modulating host response to SARS-Cov-2 across populations.

Functional studies of key genes identified are needed to identify mechanisms through which these genes influence COVID-19 related hospitalization. Additionally, while well-powered molecular genetic datasets in diverse populations often lag behind European-ancestry counter parts, massive collaborative science efforts such as the HGI-19 continue to accumulate new datasets to address this discrepancy. Leveraging population eQTL/spQTL/pQTL data with ancestry-matched COVID-19 GWAS will be crucial in identifying and understanding mechanisms underlying COVID-19 related hospitalization. In conclusion, our work raises specific hypotheses relating host genetic variation to symptom and lab-trait profiles thereby focusing efforts for future drug repurposing and therapeutic discovery research.

## METHODS AND MATERIALS

### COVID19-HGI genome-wide association summary statistics

We downloaded GWAS summary statistics for severe COVID-19 outcomes meta-analyzed across 21 studies (hospitalized N=7,885; population N=961,804). A detailed description of the contributing studies, meta analyses, and primary GWAS results for several COVID-19-related phenotypes are presented at https://www.covid19hg.org/results/, specifically the Freeze 4-October 2020 results File: COVID19_HGI_B2_ALL_leave_23andme_20201020.txt.gz. These summary statistics do not include 23&Me cohort results, and their sample size was removed from final sample reported. Genome-wide association statistics consisted of inverse-variance meta-analyzed log-odds ratios and their standard errors to compute a final Wald statistic and p-value. Most of the individuals of the contributing studies to meta-analysis genomic study were of European descent (93%). We performed strict quality control on GWAS data, by filtering statistics at palindromic variants, harmonizing variants with GTEx v8 European-panel genotypes (*29*). Our quality control procedure resulted in a final count of 10,340,768 autosomal genetic variants with summary statistics.

### TWAS and spTWAS using models of predicted gene and alternative splicing expression

To perform TWAS and spTWAS we leveraged pre-trained prediction models fitted in GTEx v8 data (49 tissues, N=838) using the fine-mapping software DAP-G with a biologically-informed prior, Multivariate Adaptive Shrinkage in R (MASHR). For detailed information regarding molecular, genetic, and phenotypic data in GTEx v8, please see (*29*). Prediction models for each tissue were integrated with COVID19-HGI GWAS data using the software S-PrediXcan (*30*). In total, we tested 655,563 and 1,728,429 models of total expression and alternative splicing, respectively for association with severe COVID19 outcomes, however, owing to the significant amount of correlation across tissues, we used a per-tissue Bonferroni correction threshold in our multi-tissue analyses (see **Table S1**). To combine association statistics across all tissues while adjusting for tissue–tissue correlation, we used S-MultiXcan (*31*). For 22,206 genes, we also performed a joint multi-tissue approach using S-MultiXcan which accounts for tissue correlation and boosts statistical power. Here, we applied a single Bonferroni correction of 0.05 / 22,206.

### PWAS using models of predicted expression

To perform a protein-wide association study (PWAS) using predicted protein expression, we fitted predictive models using genetic and plasma proteins from European-ancestry individuals in the INTERVAL study (N=3,301) (*10*). We performed quality control on genotype data and kept only bi-allelic SNPs with MAF ≧0.01, HWE p > 5e-5, imputation quality INFO > 0.6 and were annotated in HapMap3. Plasma proteins had undergone strict quality control and adjustment in the original study (*10*). We fit predictive 3,222 predictive models for 3,170 proteins using genotypes within 1Mb flanking the gene body (i.e., ±500kb gene start and stop). For measured proteins consisting of multiple monomers (i.e., dimer, trimer, etc.), we fit multiple predictors, each restricted contributing gene’s region. We included the following covariates into all downstream models of protein abundance: age, sex, duration of blood processing, the top 3 genotyping PCs, contributing cohort, and top 4 protein PCs). To reduce the number of tests and increase statistical power, we restricted to genes whose protein levels exhibited evidence of genetic control by testing for non-zero cis-heritability (p-value < 0.05) using GCTA. Our final set of local/cis-based predictors resulted in 1,031 models of protein with significant cis-SNP heritability (p-value < 0.05). We fit penalized linear models using SuSiE (*32*) and performed downstream PWAS using the tool FUSION (*33*) with the same quality controlled genome-wide association statistics as above.

### Allelic Specific Expression

To determine the allelic effect of GWAS SNPs (p-value < 5e-5) on identified susceptibility genes, we used haplotype-level ASE data with WASP filtering (*34*) from the GTEx v8, containing 15,253 samples spanning over 49 human tissues and 838 individuals (*29, 35*). We used haplotype-aggregated allelic expression generated by phASER (*36*). To assess cis-acting regulatory effect of expression imbalance between the alleles in heterozygous individuals, we compared allelic imbalance between the individuals homozygous and heterozygous for each SNP. All individuals with minimum coverage of 8 reads (with one pseudocount added) were included. Allelic imbalance was quantified as the log ratio between the two allelic counts or log allelic Fold Change (log aFC) (*37*) and to ensure robustness to rare variant effects and phasing errors the absolute value of log aFCs are compared, using a one-sided ranksum test. We used a gene-level Bonferonni correction for the total number of genes tested (p-value < 0.05 / 21).

### EHR-based PheWAS

To better understand the phenotypic consequences of dysregulated mRNA expression across our genes of interest, we performed a Phenome-Wide Association Study (PheWAS) (*38*) including patients in the Vanderbilt EHR and linked biobank, BioVU. Phenotypes in BioVU are represented as phecodes, which are assigned as a dichotomous trait and are a hierarchical clustering of the International Classification of Diseases (ICD9/ICD10) codes. For each phenotype we required a minimum number of 100 cases for inclusion in our PheWAS analyses, which resulted in testing 1,404 phecodes in 70,439 individuals of European ancestry and 740 phecodes in 15,174 individuals of African ancestry. We used the PheWAS package in R to perform logistic regressions to identify the phecodes that are significantly associated with imputed gene expression after adjusting for sex, age, and the top ten principal components from genetic data to control for population stratification (Denny et al. 2010, 2013). We corrected for the number of tests (i.e. 0.05/1404 = 3.56e-05) to determine statistical significance.

### EHR Biomarker LabWAS

The Lab-Wide Association Scan (LabWAS) (*39*) allows us to screen clinical lab tests from the Vanderbilt University Medical Center EHR. For each gene identified in the TWAS analyses, we tested the association between its predicted gene expression and all clinical labs. We applied the QualityLab cleaning pipeline (*39*) with settings to yield median age-adjusted (residual taken after regressing the cubic splines of age with 4 knots) inverse normal quantile transformed lab values (to control for skewness and non-normality). We screened across all labs with measurements for at least 100 individuals, which resulted in testing 323 labs in 70,337 individuals of European ancestry and 241 labs in 15,123 individuals of African ancestry. The lab tests are divided into 12 sub-categories; blood, metabolic, endocrine, kidney, immune, liver, urinary, OB/gyn, toxicology, cardiovascular and cancer. Our analyses included the covariates age, sex, and top ten principal components from genetic data to adjust for genetic ancestry. We used a Bonferroni-corrected threshold accounting for the number of labs present in the associations tested (i.e. 0.05/323 = 1.55e-04).

### PheWAS and LabWAS category enrichment analyses

We tested for enrichment of association signals across the clinical categories of phenotypes (laboratories) in PheWAS (LabWAS) in two ways. First, we performed a hypergeometric test using phenotypes/labs that were labelled as significant/not-significant per category. Second, we performed a relaxed test that considers for the average magnitude of association signal in a given category. Here, we computed the mean *χ*^2^ association statistic per phenotype (laboratory) category and bootstrapped its standard error using 2000 bootstraps. Our enrichment (depletion) statistics for a phenotype (laboratory) category were the difference between its mean *χ*^2^ from 1 (the expected *χ*^2^ under the null) divided by the bootstrapped standard error. We used a Bonferroni adjusted p-value < 0.05 / k to determine enrichment or depletion for either aproach, where k=17 for PheWAS category enrichment tests and k=12 for LabWAS category enrichment tests.

### Cross-ancestry gene effects

We compared effect-sizes of predicted expression on phenotypes and laboratories estimated in European-ancestry patients from the BioVU with estimates obtained from of African-ancestry records. To do so, we performed a weighted linear regression, 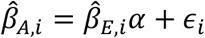, where 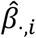., *t* are the effect-sizes estimated for the ith gene/phenotype or gene/laboratory pair in African-(A) or European-ancestry (E) patients, *α* is the cross-ancestry relationship, and 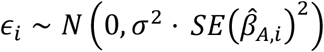 is the gene/phenotype-specific (gene/laboratory) noise parameterized by the overall variance *σ*^2^ and squared standard-error around the African-ancestry-based estimate. We report estimates of 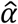 and its 95 confidence intervals assuming normality.

### Cross-ancestry SNP effects

We investigated BioVU phenotypes and laboratory measures significantly associated with TWAS-associated loci for heterogeneous effects in the trans-ethnic Pan-UK Biobank (Pan-UKB). The Pan-UKB represents a multi-ancestry analysis of 7,221 phenotypes in six continental ancestry groups: African (AFR N=6,636), Admixed American (AMR N=980), Centra/South Asian (CSA N=8,876), East Asian (EAS N=2,709), European (EUR N=420,531), and Middle Eastern (MID N=1,599). The Pan-UKB consists of 16,119 genome-wide association studies of biological assays, health status, behavioral information, and lifestyle factors. We clumped SNPs in PLINK using eQTL/sQTL p-values (q-value ≦ 0.05) reported in GTEx v8 and limited pair-wise SNP correlations to r^2^=0.1 over 250kb windows. Per-tissue clumping resulted in 27 SNPs (*ABO, IFNAR2, CCR1*, and *SLC6A20*) that were tested with respect to 1,571 Pan-UKB phenotypes from the same trait domains detected by PheWAS in BioVU (e.g., circulatory system, digestive disorder, neurological). We used linear models to test for consistency of EUR effect estimates at FDR significant and high confidence SNPs with those estimated in AFR, AMR, CSA, EAS (Pan-UKB and Japan Biobank), and MID populations. Additionally, we used Biobank Japan to verify associations between EUR and EAS-based SNP effects in the Pan-UKB due to increased sample size in the latter. Biobank Japan consists of genetic data for over 200,000 participants and ∼120 disease states and quantitative measures (cell type percentage, body mass index, etc.).

## Data Availability

All data is provided in the supplementary files and full results are made available via dropbox

## Acknowledgement

We thank the COVID-19 Host Genetics Initiative (https://www.covid19hg.org/acknowledgements/) for providing open access to genetic association data, and all the studies comprising of the meta-analysis (https://www.covid19hg.org/publications/).

## Author contributions

GAP, KS, and NAM contributed to study design, writing, and editing the manuscript. GAP, KS, TMF, FRW, NE, and ZL contributed to the study design and analysis. All the authors provided critical feedback on the manuscript, study design and expertise of multiple methods contributed to the manuscript. NAM conceptualized and headed the TWAS-working group within the COVID-19 HGI, supervised drafting, and finalized the manuscript with LKD, BP, and RP.

## Data and materials availability

The data is available in the attached Supplementary files, and full results can be accessed from https://www.dropbox.com/sh/gauoyuvmqak77fc/AADhZJT2eHNUUhTumqzM_I5oa?dl=0

## CAPTIONS AND LEGEND FOR SUPPLEMENTARY MATERIAL

### List of Supplementary Materials

#### Supplementary Materials

##### Supplementary Tables: Supplementary_TablesS1-S20.xlsx

Table S1. Significant results for the TWAS from multi-tissue approach using S-MetaXcan.

Table S2. Number of TWAS tests per tissue using GTEx v8 data.

Table S3. All TWAS results. Please see dropbox link.

Table S4: Results of expression TWAS using meta-analyzed across tissue approach using SMultiXcan, significant results are highlighted in yellow.

Table S5: Significance analysis of allelic imbalance of genes in their respective tissues using leading GWAS variants (or closest proxy) within 1Mb region.

Table S6: Significant results for the spTWAS from multi-tissue approach using S-MetaXcan.

Table S7. Number of spTWAS tests per tissue using GTEx v8 data.

Table S8. All spTWAS results. Please see dropbox link.

Table S9. Results of splicing TWAS using meta-analyzed across tissue approach using SMultiXcan, significant results are highlighted in yellow. Results shown here are p<0.1.

Table S10: Comparing effect size estimates of genes that were significant using both eTWAS and spTWAS

Table S11: All results for proteome/protein transcriptome-wide association study. The significant genes are highlighted in yellow.

Table S12. Significant results for PheWAS-phenome-wide association study using the genetically-predicted gene expression.

Table S13: Enrichment of phenotype domains for the significant phenotypes

Table S14: Enrichment of phenotype domains for all phenotypes

Table S15. Significant results for LabWAS-Laboratory-test-wide association study using the genetically-predicted gene expression.

Table S16: Enrichment of lab domains for the significant lab-phenotypes

Table S17: Enrichment of lab domains for the all lab-phenotypes

Table S18: PheWAS of e/spQTL SNPs for the gene/tissues identified from GTEx-based Phe/LabWAS in the Pan-UK biobank

Table S19: PheWAS of e/spQTL SNPs for the gene/tissues identified from GTEx-based Phe/LabWAS in the Biobank Japan and its comparison with Pan-UK biobank

Table S20: Results from Linear Models of Non-European Effect Sizes in Relation To European Effect Sizes

##### Supplementary Figures: Supplementary_FigS1-S4.docx

Figure S1: Manhattan plot of the meta-analyzed tissue TWAS of gene expression using SMultiXcan.

Figure S2: Allelic imbalance detected for gene/tissue pairs identified from TWAS, using leading GWAS variants (or closest proxy)

Figure S3: Manhattan plot of the meta-analyzed tissue TWAS of splicing expression using SmultiXcan.

Figure S4: Venn diagram showing overlap of significant genes identified from TWAS, spTWAS and PWAS.

## REFERENCES

1. R. Lu et al., Genomic characterisation and epidemiology of 2019 novel coronavirus: implications for virus origins and receptor binding. Lancet. 395, 565–574 (2020).

2. S. Richardson et al., Presenting Characteristics, Comorbidities, and Outcomes Among 5700 Patients Hospitalized With COVID-19 in the New York City Area. JAMA. 323, 2052–2059 (2020).

3. L. Mao et al., Neurologic manifestations of hospitalized patients with coronavirus disease 2019 in Wuhan, China. JAMA Neurol. (2020), doi:10.1001/jamaneurol.2020.1127.

4. M. Hoffmann et al., SARS-CoV-2 Cell Entry Depends on ACE2 and TMPRSS2 and Is Blocked by a Clinically Proven Protease Inhibitor. Cell. 181, 271–280.e8 (2020).

5. E. Pairo-Castineira et al., Genetic mechanisms of critical illness in Covid-19. medRxiv (2020), doi:10.1101/2020.09.24.20200048.

6. COVID-19 Host Genetics Initiative, The COVID-19 Host Genetics Initiative, a global initiative to elucidate the role of host genetic factors in susceptibility and severity of the SARS-CoV-2 virus pandemic. Eur. J. Hum. Genet. 28, 715–718 (2020).

7. G. H. L. Roberts et al., AncestryDNA COVID-19 Host Genetic Study Identifies Three Novel Loci. medRxiv (2020), doi:10.1101/2020.10.06.20205864.

8. J. F. Shelton et al., Trans-ethnic analysis reveals genetic and non-genetic associations with COVID-19 susceptibility and severity. medRxiv (2020), doi:10.1101/2020.09.04.20188318.

9. Severe Covid-GWAS Group, Genomewide association study of severe Covid-19 with respiratory failure. New England Journal of … (2020).

10. B. B. Sun et al., Genomic atlas of the human plasma proteome. Nature. 558, 73–79 (2018).

11. S. Tal, G. Spectre, R. Kornowski, L. Perl, Venous Thromboembolism Complicated with COVID-19: What Do We Know So Far? Acta Haematol. 143, 417–424 (2020).

12. G. H. Frydman, E. W. Boyer, R. M. Nazarian, E. M. Van Cott, G. Piazza, Coagulation Status and Venous Thromboembolism Risk in African Americans: A Potential Risk Factor in COVID-19. Clin. Appl. Thromb. Hemost. 26, 1076029620943671 (2020).

13. A. Porfidia, A. Santoliquido, G. Cammá, E. Porceddu, R. Pola, Incidence of Deep Vein Thrombosis among non-ICU Patients Hospitalized for COVID-19 Despite Pharmacological Thromboprophylaxis. J. Thromb. Haemost. (2020), doi:10.1111/jth.15089.

14. F. A. Klok et al., Incidence of thrombotic complications in critically ill ICU patients with COVID-19. Thromb. Res. 191, 145–147 (2020).

15. F. Grillet, J. Behr, P. Calame, S. Aubry, E. Delabrousse, Acute Pulmonary Embolism Associated with COVID-19 Pneumonia Detected with Pulmonary CT Angiography. Radiology. 296, E186–E188 (2020).

16. G. Lu, J. Wang, Dynamic changes in routine blood parameters of a severe COVID-19 case. Clin. Chim. Acta. 508, 98–102 (2020).

17. S. M. Rezende, W. M. Lijfering, F. R. Rosendaal, S. C. Cannegieter, Hematologic variables and venous thrombosis: red cell distribution width and blood monocyte count are associated with an increased risk. Haematologica. 99, 194–200 (2014).

18. F. H. Algahtani, R. Stuckey, High factor VIII levels and arterial thrombosis: illustrative case and literature review. Ther. Adv. Hematol. 10, 2040620719886685 (2019).

19. M. Sabater-Lleal et al., Genome-Wide Association Transethnic Meta-Analyses Identifies Novel Associations Regulating Coagulation Factor VIII and von Willebrand Factor Plasma Levels. Circulation. 139, 620–635 (2019).

20. K. M. Shepardson et al., IFNAR2 Is Required for Anti-influenza Immunity and Alters Susceptibility to Post-influenza Bacterial Superinfections. Front. Immunol. 9, 2589 (2018).

21. L. Prokunina-Olsson et al., COVID-19 and emerging viral infections: The case for interferon lambda. J. Exp. Med. 217 (2020), doi:10.1084/jem.20200653.

22. M. Sa Ribero, N. Jouvenet, M. Dreux, S. Nisole, Interplay between SARS-CoV-2 and the type I interferon response. PLoS Pathog. 16, e1008737 (2020).

23. C. G. K. Ziegler et al., SARS-CoV-2 Receptor ACE2 Is an Interferon-Stimulated Gene in Human Airway Epithelial Cells and Is Detected in Specific Cell Subsets across Tissues. Cell. 181, 1016–1035.e19 (2020).

24. Q. Zhang et al., Inborn errors of type I IFN immunity in patients with life-threatening COVID-19. Science. 370 (2020), doi:10.1126/science.abd4570.

25. P. Bastard et al., Autoantibodies against type I IFNs in patients with life-threatening COVID-19. Science. 370 (2020), doi:10.1126/science.abd4585.

26. E. Meffre, A. Iwasaki, Interferon deficiency can lead to severe COVID. Nature (2020), doi:10.1038/d41586-020-03070-1.

27. J. A. Membrilla, Í. de Lorenzo, M. Sastre, J. Díaz de Terán, Headache as a Cardinal Symptom of Coronavirus Disease 2019: A Cross-Sectional Study. Headache (2020), doi:10.1111/head.13967.

28. J. C. García-Moncó et al., Neurological reasons for consultation and hospitalization during the COVID-19 pandemic. Neurol. Sci. 41, 3031–3038 (2020).

29. GTEx Consortium, The GTEx Consortium atlas of genetic regulatory effects across human tissues. Science. 369, 1318–1330 (2020).

30. A. N. Barbeira et al., Exploring the phenotypic consequences of tissue specific gene expression variation inferred from GWAS summary statistics. Nat. Commun. 9, 1825 (2018).

31. A. N. Barbeira et al., Integrating predicted transcriptome from multiple tissues improves association detection. PLoS Genet. 15, e1007889 (2019).

32. Q. S. Wang et al., Leveraging supervised learning for functionally-informed fine-mapping of cis-eQTLs identifies an additional 20,913 putative causal eQTLs. BioRxiv (2020), doi:10.1101/2020.10.20.347294.

33. A. Gusev et al., Integrative approaches for large-scale transcriptome-wide association studies. Nat. Genet. 48, 245–252 (2016).

34. B. van de Geijn, G. McVicker, Y. Gilad, J. K. Pritchard, WASP: allele-specific software for robust molecular quantitative trait locus discovery. Nat. Methods. 12, 1061–1063 (2015).

35. S. E. Castel et al., A vast resource of allelic expression data spanning human tissues. Genome Biol. 21, 234 (2020).

36. S. E. Castel, P. Mohammadi, W. K. Chung, Y. Shen, T. Lappalainen, Rare variant phasing and haplotypic expression from RNA sequencing with phASER. Nat. Commun. 7, 12817 (2016).

37. P. Mohammadi, S. E. Castel, A. A. Brown, T. Lappalainen, Quantifying the regulatory effect size of cis-acting genetic variation using allelic fold change. Genome Res. 27, 1872–1884 (2017).

38. J. C. Denny et al., PheWAS: demonstrating the feasibility of a phenome-wide scan to discover gene-disease associations. Bioinformatics. 26, 1205–1210 (2010).

39. J. K. Dennis et al., Lab-wide association scan of polygenic scores identifies biomarkers of complex disease. medRxiv (2020), doi:10.1101/2020.01.24.20018713.

